# The Genetic Association of SP-A and SP-D Polymorphisms with Tuberculosis and Latent TB in the Pakistani Population

**DOI:** 10.64898/2026.03.04.26347180

**Authors:** Urooj Subhan, Farah Deeba, Ezza Binte Tariq, Manahil Tahir, Afrose Liaquat, Sidra Younis

## Abstract

**Background:** Tuberculosis (TB) remains a major global health concern, posing a substantial burden in Pakistan. Genetic factors play a pivotal role in individual susceptibility to TB. Surfactant protein A and surfactant protein D are essential components of the innate immune system, contributing to pulmonary host defense against *Mycobacterium tuberculosis* (MTB).

**Aim:** This study aimed to investigate the association between single nucleotide polymorphisms (SNPs) in the SP-A1gene at rs1059047 (+1101 C/T) and the SP-D gene at rs3088308 (911 T/A) and TB susceptibility in the Pakistani population.

**Method:** A case-control study was conducted, comprising 350 individuals, including 150 healthy controls, 100 TB patients, and 100 TB contacts. Genotyping was performed using tetra ARMS-PCR. GraphPad Prism v.10 was used for statistical analysis.

**Results:** Our results revealed no significant association between the SP-A1 gene polymorphism at rs1059047 (+1101 C/T) and SP-D gene polymorphism at rs3088308 (911 T/A) and TB susceptibility in the Pakistani population (p>0.5). Interestingly, concerning the rs3088308 polymorphism in the SP-D gene, a comparison between healthy controls and TB contacts indicated that the homozygous TT genotype was significantly associated with protection against LTBI (73.53% *vs*. 82.35%; p=0.00, OR=0.19, 95% CI=0.08-0.51).

## Introduction

Tuberculosis (TB) is an airborne contagious disease primarily caused by the bacterium *Mycobacterium tuberculosis* (MTB) (1). In 2024, 10.6 million people were infected with TB globally, and 1.6 million people died from TB (2). Pakistan is ranked 5^th^ among highly prevalent TB countries (3). A quarter of the world’s population is infected with MTB commonly referred as latent TB infection (LTBI) (4).

TB spreads through aerosol droplets containing MTB, expelled by active TB patients, followed by inhalation and deposition into new hosts. At this stage, host innate immunity plays a crucial role in controlling infection, leading to the internalization of MTB by alveolar macrophages. However, when these macrophages are unable to effectively inhibit or destroy the bacilli, the bacteria multiply within the intracellular environment and are subsequently released and phagocytosed by other alveolar macrophages (5). Recruitment and multiplication of new lymphocytes to the site of infection instigate a cell-mediated immune response in which a multitude of immune cells strive to confine the bacteria and curtail their further proliferation (6). During this phase, the host typically remains asymptomatic, and there is the possibility of complete elimination of MTB or their transition to a latent state within granulomas. Nonetheless, when the host immune system is compromised, the latent infection can rapidly progress to active disease, accompanied by clinical symptoms (7).

The water-soluble C-type lectins known as SP-A1 and SP-D, belonging to the collectin family, are involved in the innate immune system. SP-A1, along with SP-D, plays a crucial role in the elimination of MTB by enhancing its phagocytosis by macrophages (8, 9). SP-A1 acts as a bridge between MTB and macrophages by interacting with the glycoprotein located on the surface of MTB and mannose receptors located on the surface of alveolar macrophages, thereby enhancing the engulfment of MTB (10). Similarly, SP-D participates in the intrinsic immunological reaction. It recognizes and binds to specific carbohydrate sequences present on the surface of various microbes, facilitating their clearance by immune cells. SP-D also aids in surfactant homeostasis, modulation of inflammation, and lung defence against pathogens (11).

SP-A1 and SP-D genetic variants are associated with both chronic and acute lung diseases, as well as some non-pulmonary diseases (12-14). However, there is a lack of data on the association between SP-A1 and SP-D SNPs with TB/LTBI in the Pakistani population. Therefore, we planned to investigate the association between SP-A1 (rs1059047) 1101 T/C located at exon 3 of the SP-A1 gene and SP-D (rs3088308) 911 T/A located at exon 7 of the SP-D gene with TB susceptibility in the Pakistani population.

## Methodology

### Ethical Approval

This study was approved by the Institutional Review Board of the National University of Medical Sciences (NUMS) under reference number 06/IRB&EC/NUMS/24 (S1). Informed written consent was obtained from each participant of the study in accordance with the Helsinki Declaration of 1975 as revised in 2013 (S2).

### Study Population

In the present case-control study a total of 350 blood samples were collected from Nishtar Hospital Multan (NHM) and Raza Laboratory, Nishtar Road Multan. The samples included 100 active TB patients, 100 TB contacts (excluding blood relations), and 150 healthy controls with ages ranging from 17-72 years. The sample size was calculated using Epitool software. All the study participants were aged 16 or above. Newly diagnosed active TB patients, confirmed by GeneXpert or chest X-ray, were included in the patient group. Household contacts with index cases of pulmonary TB within the last 6 months and asymptomatic were included in the contacts group. Asymptomatic individuals with no history of contact with TB patients were recruited as healthy individuals. All the participants gave written informed consent to participate in the study. Individuals with known anaemia, HIV infection, pregnancy, or breastfeeding were excluded from the study (S3).

### DNA Extraction

DNA was extracted using the phenol-chloroform method (15). The quality and purity of extracted DNA were checked by Nanodrop.

### Tetra ARMS PCR

Primers were designed using Primer 1 software. The tetra-arms PCR using 2 sets of primers was used to selectively amplify the target region of interest. Gradient PCR was used for the optimization of the annealing temperature. PCR products were separated on 2% agarose gel and visualized by Gel Doc XR+ Gel Documentation System. Nucleotide sequences of primers with their properties and PCR cycle details are given in Table 1 and Table 2.

**Table 1:**
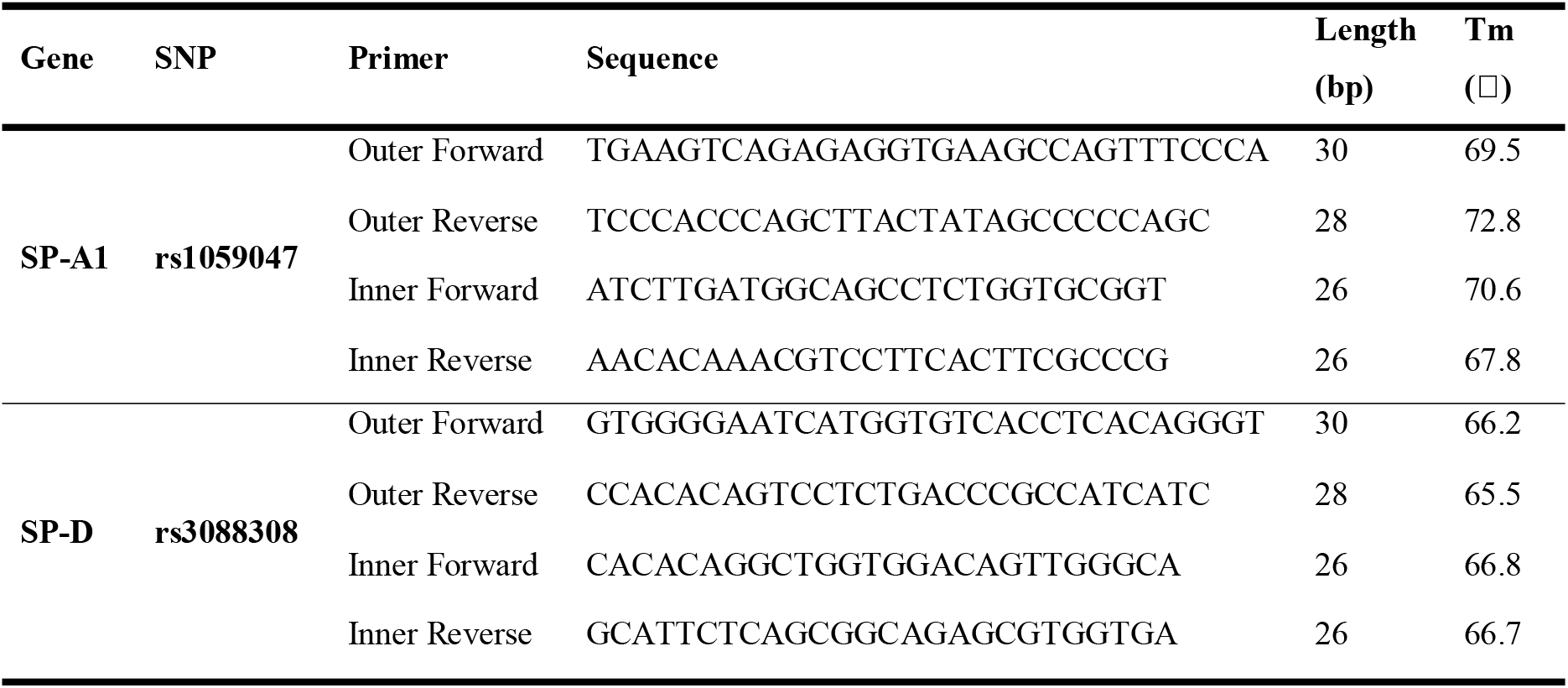
Sequence and properties of primers.

**Table 2:** PCR conditions for all mutations.

| SNP | PCR cycle details |  |  |  | Final extension |
| --- | --- | --- | --- | --- | --- |
|  | Initial denaturation | Denaturation | Annealing (35 cycles) | Extension |  |
| **rs1059047** | 95 °C for 5 min | 95 °C for 1 min | 69 °C for 1 min | 72 °C for 1 min | 72 °C for 10 min |
| **Rs3088308** | 95 °C for 5 min | 95 °C for 1 min | 73 °C for 1 min | 72 °C for 1 min | 72 °C for 10 min |

### Statistical analysis

GraphPad Prism 10 software was used to statistically analyze the data. For each SNP, Hardy-Weinberg Equilibrium (HWE) was calculated using the chi-square test. Continuous data was analyzed using independent sample t-test. Genotype frequencies were compared using the chi-square test.

## Results

The mean age of patients, contacts, and healthy controls was 40.85 ± 16.19 years, 37.39 ± 10.62 years, and 40.85 ± 16.19 years, respectively. The number of females *vs*. males in the study subjects was as follows: TB patients: 57 *vs*. 43, Contacts: 49 *vs*. 51, and Controls: 76 *vs*. 74, respectively.

### Association of SP-A1 (rs1059047) with active TB and TC

ARMS PCR was used to analyse the genotype pattern of SNP rs1059047 with TB and LTBI. PCR products were resolved on 2% agarose gel and genotypes were assessed by band distribution pattern (Figure 1). We obtained two bands of 663 and 264 bp for TT (homozygous wild type), two bands of 663 and 450 bp for CC (homozygous mutant type), and three bands for 663, 450 and 264 bp for CT (heterozygous mutant type).

**Figure 1:**
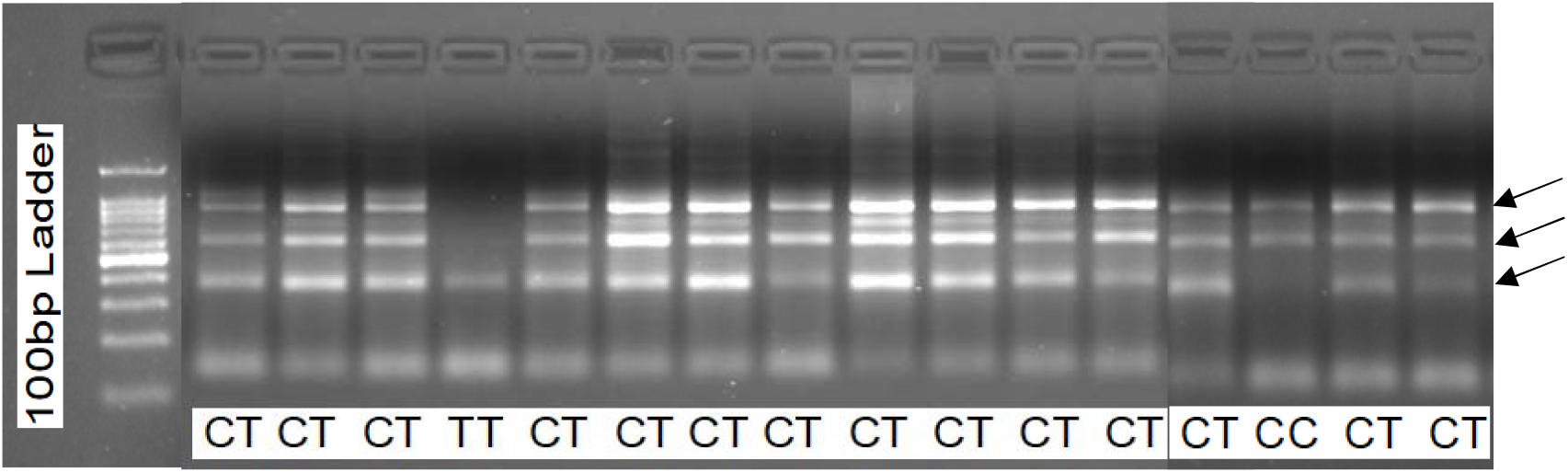
Agarose gel picture representing genotype pattern for rs1059047. Lane 1 represents 100 bp DNA ladder. The other lanes refer to the genotype pattern. Product sizes are characterized as 663+450 bp for CC, 663+264 bp for TT, and 663+450+264 bp for CT.

When the genotypes were compared between healthy controls and TB patients in a co-dominant model, a statistically insignificant p-value was obtained (p=0.57, OR=0.65, 95%CI=0.03-12.52). The frequency of CT was similar in healthy controls (91.72%) and TB patients (93.68%). The genotype frequencies of TT and CC in healthy controls *vs*. TB patients had a minor difference (Healthy controls *vs*. TB Patients; TT: 0.69% *vs*. 1.05% and CC: 7.59% *vs*. 5.26%,). Similar genotype comparisons in the dominant model, recessive model, and additive model gave statistically non-significant p-values (Dominant Model: p=0.76, OR=0.65, 95%CI=0.03-12.52; recessive model: p=0.48, OR=1.47, 95%CI=0.48-3.93 and additive model: p=0.26, OR=0.95, 95%CI=0.65-1.37) (Table 3a).

**Table 3a.** Genotype comparison for SP-A1 rs1059047 between healthy controls and TB patients.

| rs1059047 | Genotypes | HC<br>n (%) | TP<br>n (%) | OR<br>(95% CI) | p<br>value | $\chi^2$<br>Value |
| --- | --- | --- | --- | --- | --- | --- |
|  |  | 145 (100) | 95 (100) |  |  |  |
| **Codominant model** | TT | 1 (0.69) | 1 (1.05) | 0.65 (0.03-12.52) | 0.74 | 0.57 |
|  | CT | 133 (91.72) | 89 (93.68) |  |  |  |
|  | CC | 11 (7.59) | 5 (5.26) |  |  |  |
| **Dominant Model** | TT | 1 (0.69) | 1 (1.05) | 0.65 (0.03-12.52) | 0.76 | 0.09 |
|  | CT + CC | 144 (99.31) | 94 (98.95) |  |  |  |
| **Recessive Model** | CC | 11 (7.59) | 5 (5.26) | 1.47 (0.48-3.93) | 0.48 | 0.49 |
|  | CT + TT | 134 (92.41) | 90 (94.74) |  |  |  |
| **Additive model** | T | 134 (46.37) | 91 (47.89) | 0.95 (0.65-1.37) | 0.26 | 0.07 |
|  | C | 155 (53.63) | 99 (52.38) |  |  |  |

$\chi^2$ : chi square, OR: odds ratio CI: confidence interval, HC: Healthy Control, TP: TB Patient.

We have also compared genotypes between healthy controls and TB contacts in co-dominant, dominant, recessive, and additive models. We found a statistically insignificant p-value in the co-dominant model (p=0.25, OR=0.68, 95% CI=0.03-13.18). The frequency of the TT genotype was similar in healthy controls *vs*. TB contacts (0.69% *vs*. 1.00%), whereas the frequency of the CT genotype was higher in healthy controls compared to TB contacts (91.72% *vs*. 85.00%). Similarly, the frequency of the CC genotype was different but statistically non-significant among study groups (Healthy controls: 7.59% and TB contacts: 14.00%). Furthermore, genotype comparison in the dominant model, recessive, and additive model produced statistically insignificant p-values (Dominant model; p=0.79, OR=0.68 95% CI=0.03-13.18; Recessive Model; p=0.10, OR=0.50, 95%CI=0.23-1.14 and additive model; p=0.53, OR=1.12, 95%CI=0.78-1.61) (Table 3b).

**Table 3b.** Genotype comparison for SP-A1 rs1059047 between healthy controls and TB contacts.

| rs1059047 | Genotypes | HC | TC | OR<br>(95% CI) | p<br>value | $\chi^2$<br>Value |
| --- | --- | --- | --- | --- | --- | --- |
|  |  | n (%) | n (%) |  |  |  |
|  |  | **145 (100)** | **100 (100)** |  |  |  |
| **Codominant model** | TT | 1 (0.69) | 1 (1.00) | 0.68 (0.03-13.18) | 0.25 | 2.75 |
|  | CT | 133 (91.72) | 76 (85.00) |  |  |  |
|  | CC | 11 (7.59) | 14 (14.00) |  |  |  |
| **Dominant Model** | TT | 1 (0.69) | 1 (1.00) | 0.68 (0.03-13.18) | 0.79 | 0.07 |
|  | CT + CC | 144 (99.31) | 99 (99.00) |  |  |  |
| **Recessive Model** | CC | 11 (7.59) | 14 (14.00) | 0.50 (0.23-1.14) | 0.10 | 2.65 |
|  | CT + TT | 134 (92.41) | 77 (86.00) |  |  |  |
| **Additive model** | T | 134 (46.37) | 78 (43.50) | 1.12 (0.78-1.61) | 0.53 | 0.39 |
|  | C | 155 (53.63) | 102 (56.50) |  |  |  |

$\chi^2$ : chi square, OR: odds ratio CI: confidence interval, HC: Healthy Control, TC: TB Contact.

### Association of SP-D (rs3088308) with active TB and TC

ARMS PCR was used to analyse the genotype pattern of SNP rs1059047 with TB and TC. PCR products were resolved on 2% agarose gel and genotypes were assessed by band distribution pattern (Figure 2). We obtained two bands of 515 and 347 bp for TT (homozygous wild type), two bands of 515 and 217 bp for AA (homozygous mutant type), and three bands of 515, 347 and 217 bp for AT (heterozygous mutant type).

**Figure 2:**
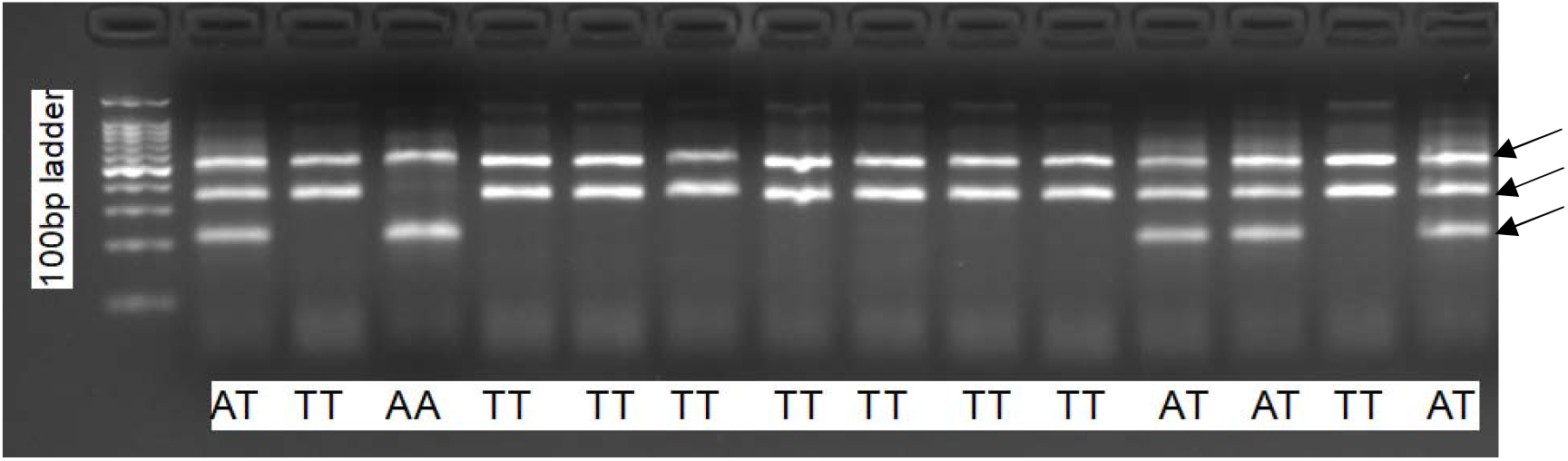
Agarose gel picture representing genotype pattern for rs3088308. Lane 1 represents 100 bp DNA ladder. The other lanes refer to genotype pattern. Product sizes are characterized as 515+347 bp for TT, 515+217 bp for AA, and 515+347+217 bp for AT.

We have also compared the genotypes in healthy controls *vs*. TB patients and healthy controls *vs*. TB contacts for SP-D rs3088308 T/A polymorphism. When the genotypes were compared between healthy controls and TB patients in a co-dominant model, a statistically insignificant p-value was obtained (p=0.74, OR=0.79, 95%CI=0.43-1.45). The difference in the frequency of TT, AT, and AA genotypes between healthy controls and TB patients was not significant (TT: 73.52% *vs*. 77.77%, AT: 19.85% *vs*. 17.17%, and AA: 6.61% *vs*. 5.05%). Similarly, we did not find statistically significant p-values for the genotype comparisons in the dominant model, recessive model, and additive model (Dominant: p=0.45, OR=0.79, 95%CI=0.43-1.45; Recessive: p=0.61, OR=1.33, 95%CI=0.44-3.64 and Additive: p=0.38, OR=0.79, 95%CI=0.47-1.31) (Table 4a).

**Table 4a:** Genotype comparison for SP-D rs3088308 between healthy controls and TB patients.

| rs3088308 | Genotypes | HC<br>n (%) | TP<br>n (%) | OR<br>(95% CI) | p<br>value | $\chi^2$<br>Value |
| --- | --- | --- | --- | --- | --- | --- |
|  |  | **136 (100)** | **99 (100)** |  |  |  |
| **Codominant model** | TT | 100 (73.53) | 77 (77.78) | 0.79 (0.43-1.45) | 0.74 | 0.59 |
|  | AT | 27 (19.85) | 17 (17.17) |  |  |  |
|  | AA | 9 (6.62) | 5 (5.50) |  |  |  |
| **Dominant Model** | TT | 100 (73.53) | 77 (77.78) | 0.79 (0.43-1.45) | 0.45 | 0.55 |
|  | AT + AA | 36 (26.47) | 22 (22.22) |  |  |  |
| **Recessive Model** | AA | 9 (6.62) | 5 (5.50) | 1.33 (0.44-3.64) | 0.61 | 0.25 |
|  | AT + TT | 127 (93.38) | 94 (94.95) |  |  |  |
| **Additive model** | T | 227 (83.46) | 171 (86.36) | 0.79 (0.47-1.31) | 0.38 | 0.74 |
|  | A | 45 (16.54) | 27 (13.64) |  |  |  |

$\chi^2$ : chi square, OR: odds ratio CI: confidence interval, HC: Healthy Control, TP: TB Patient.

Interestingly, when the genotypes were compared between healthy controls and TB contacts in the co-dominant model, a statistically significant p-value was obtained (p=0.00, OR=0.19, 95% CI=0.08-0.51). The frequency of genotype TT was higher in TB contacts than in healthy controls (82.35% *vs*. 73.53%). The frequency of AT and AA showed a minor difference among study groups (Healthy controls *vs*. TB contacts: AT: 19.85% *vs*. 16.47% and AA: 6.62% *vs*. 1.72%). Genotype comparison in the dominant model and additive model produced statistically significant p-values, while in the recessive model, a statistically insignificant p-value was obtained (Dominant model: p<0.00, OR=0.19 95% CI=0.08-0.51; Additive model: p<0.00, OR=0.16, 95%CI=0.06-41; Recessive model: p=0.15, OR=4.03, 95%CI=0.63-45.06)(Table 4b). The distribution of genotype frequencies of the studied polymorphism in study groups are diagrammatically represented in Figure 3.

**Table 4b.** Genotype comparison for SP-D rs3088308 between healthy controls and TB contacts.

| rs3088308 | Genotypes | HC<br>n (%) | TC<br>n (%) | OR<br>(95% CI) | p<br>value | $\chi^2$<br>Value |
| --- | --- | --- | --- | --- | --- | --- |
|  |  | **136 (100)** | **85 (100)** |  |  |  |
| **Codominant** | TT | 100 (73.53) | 70 (82.35) | 0.19 (0.08-0.51) | **0.00** | 10.22 |
| **model** | AT | 27 (19.85) | 14 (16.47) |  |  |  |
|  | AA | 9 (6.62) | 1 (1.72) |  |  |  |
| **Dominant Model** | TT | 100 (73.53) | 70 (93.33) | 0.19 (0.08-0.51) | **0.00** | 12.11 |
|  | AT + AA | 36 (26.47) | 15 (6.67) |  |  |  |
| **Recessive Model** | AA | 9 (6.62) | 1 (1.72) | 4.03 (0.63-45.06) | 0.15 | 1.92 |
|  | AT + TT | 127 (93.38) | 84 (98.28) |  |  |  |
| **Additive model** | T | 227 (83.46) | 154 (96.84) | 0.16 (0.06-0.41) | **0.00** | 7.57 |
|  | A | 45 (16.54) | 16 (3.11) |  |  |  |

$\chi^2$ : chi square, OR: odds ratio CI: confidence interval, HC: Healthy Control, TC: TB Contact. The significant *p* values are shown in bold.

**Figure 3:**
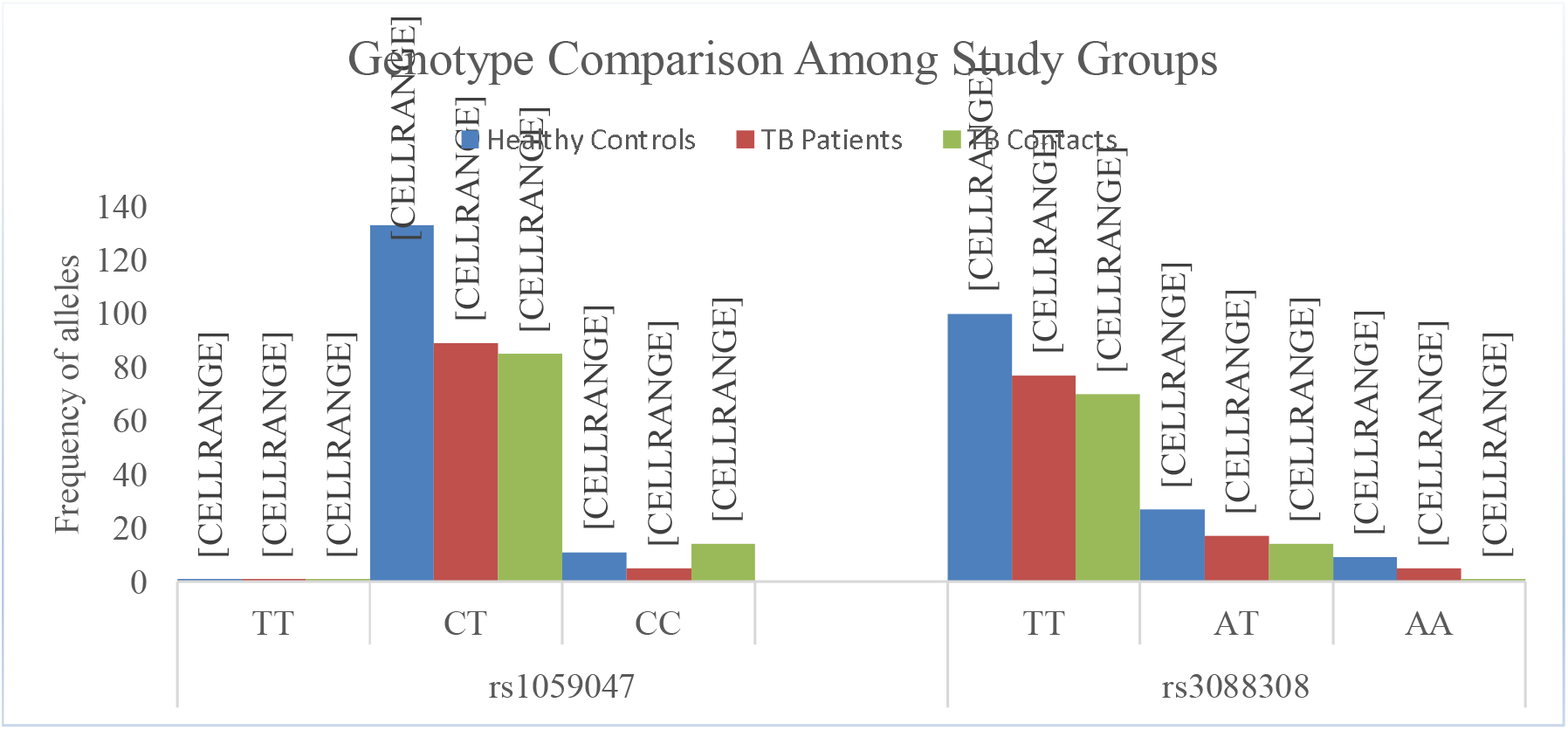
Graph representing genotype pattern among the study group.

## Discussion

The human innate immunity genes SP-A1 and SP-A2 genes are important in TB pathogenesis because they interact with MTB receptors and regulate its clearance. The polymorphisms rs1059047 and rs3088308 regulate SP-A1 and SP-D gene expression. However, their role in LTBI and TB has not been studied yet. Therefore, we conducted this study to find out the association of rs1059047 (+1101C/T) and rs3088308 (911T/A) polymorphisms in TB patients and TC from the Pakistani population.

The results of the present study revealed no significant association between the rs1059047 SP-A1 (+1101 C/T) genotype/allele frequencies and TB susceptibility in the Pakistani population. The SNP rs1059047 (missense variant) is located at exon 3, introducing a nucleotide change from T>C and amino acid change from valine to alanine at position 19. However, the substitution can affect the local interactions within the protein, lower the protein serum level, and alter its secondary structure. These results are consistent with those described in previous case-control studies conducted by Malik et al. (2006) aimed at investigating genetic variants in the SP-A1 and SP-A2 genes and their association with susceptibility to TB in Ethiopia. In their study, nine SNPs located in the exonic region of SP-A1 and SP-A2 were examined. They found that SP-A1 gene polymorphism at rs1050947 is not associated with TB, while SNPs aa62 and aa219 are significantly associated with an increased susceptibility to TB (16). Similarly, in the Chinese Han population, Yang et al. reported that aa91-G (Pro-Ala) and aa9-C are associated with TB susceptibility, while rs1059047 aa19 variant is not associated (17).

The results of the current study showed that for the comparison of SP-D gene polymorphism at rs3088308 T>A between healthy controls and TB contacts, the homozygous TT genotype was significantly associated with protection against LTBI. The SNP rs3088308 (missense variant) is located at exon 7, introducing a nucleotide change from A>T 4 and amino acid change from threonine to serine at position 270, leading to increased serum SP-D levels (18). The carbohydrate recognition domain (CRD) of SP-D interacts with terminal monosyl units (mannose cap) present on the surface of MTB, comprising lipoglycan and lipoarabinomannan, located on the surface of MTB. Upon interaction, it stimulates agglutination of MTB bacilli, ultimately leading to decreased bacterial uptake by alveolar macrophages leading to reduced MTB growth in monocytes-derived macrophages. Through altering the host cell function, such as the production and release of cytokines, reactive oxygen species (ROS) formation, or modulation of phagocytic interaction between microbes and host cell, agglutination enhances the mucociliary clearance of bacteria and viruses (19).

This investigation explored a population-specific association between SP-A1 and SP-D polymorphisms and TB/LTBI in a Pakistani cohort. While the sample size was modest and therefore not intended to provide definitive evidence, it was calculated using appropriate software and offers preliminary insight into the potential relevance of these polymorphisms. LTBI classification was based on contact history rather than IGRA or QuantiFERON testing. ARMS-PCR, although cost-effective, may not detect low-frequency variants. Consequently, these findings should be interpreted as exploratory, and further studies incorporating larger cohorts, standardized LTBI diagnostics, and functional validation of the identified SNPs are warranted to clarify their biological significance in TB pathogenesis.

## Data Availability

All data produced in the present study are available upon reasonable request to the authors

## Declarations

### Ethics approval and consent to participate

This study was approved by the Institutional Review Board of the National University of Medical Sciences (NUMS) under reference number 06/IRB&EC/NUMS/24 (S1).

### Consent for Publication

Not applicable.

### Availability of Data and Materials

All data generated or analyzed during this study are included in this published article [and its supplementary information files].

### Authors’ contributions

Urooj Subhan: Data Curation, Formal Analysis, Investigation, Writing Original Draft Dr. Farah Deeba: Project Administration, Supervision, Writing-Review and Editing Ezza Binte Tariq: Data Curation, Formal Analysis, Investigation

Manahil Tahir: Data Curation Afrose Liaquat: Data Curation,

Dr. Sidra Younis: Conceptualization, Project Administration, Methodology, Supervision, Visualization Validation, Writing-Review and Editing

Software: Primer 1, OligoIDT Analyzer, Graphpad Prism,

Resources and Funding Acquisition: National University of Medical Sciences, Rawalpindi, and The Women University, Multan.

## Acknowledgements

We acknowledge the study subjects who have voluntarily provided their blood samples for this investigation.

