## Supplementary File for "The Genetic Association of SP-A and SP-D Polymorphisms with Tuberculosis and Latent TB in the Pakistani Population"

**Supplementary material**

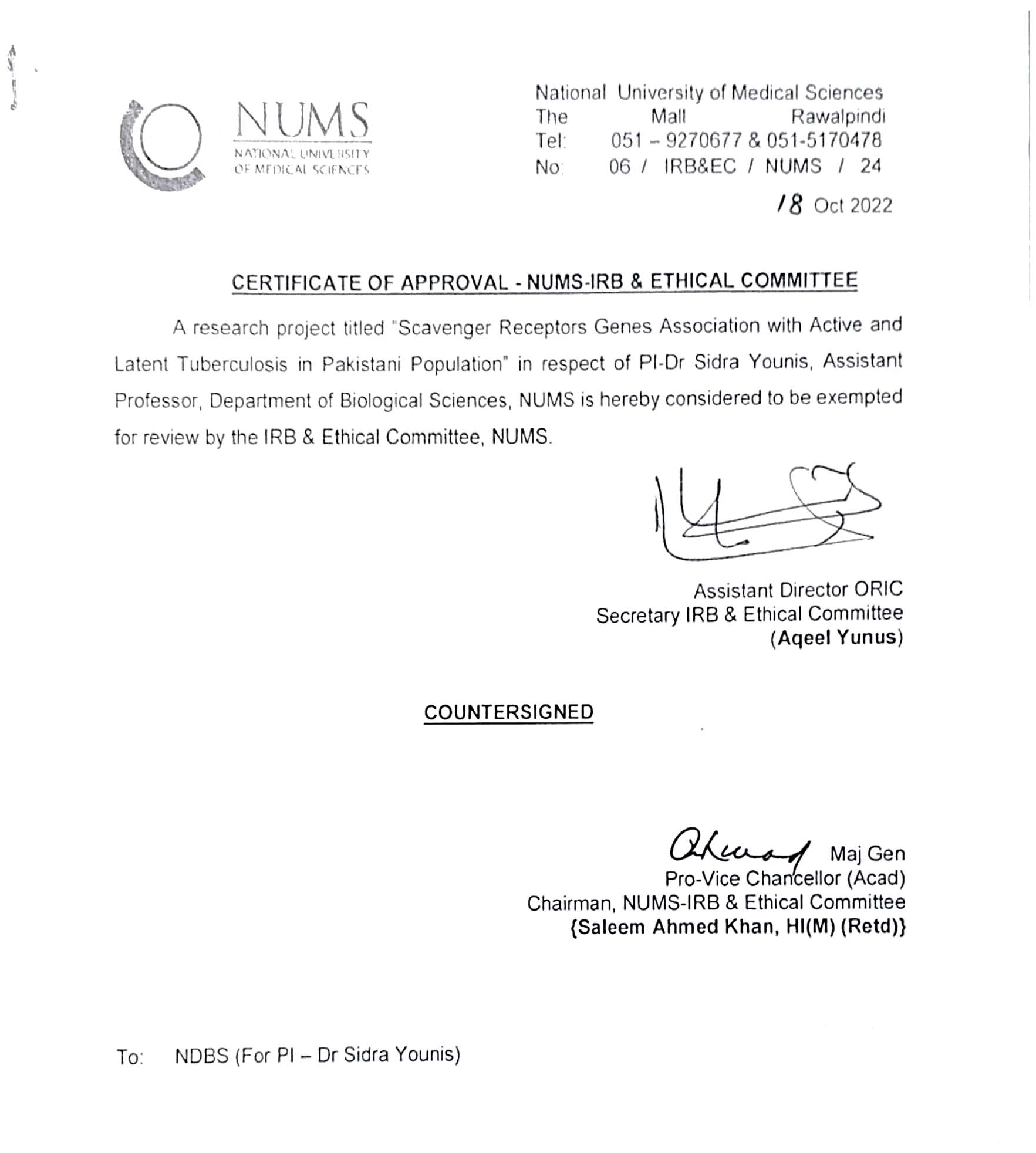
**Ethical Approval**

**Consent Form**

**Title of Study**

The Genetic Association of SP-A and SP-D Polymorphisms with Tuberculosis and Latent TB in the Pakistani Population.

**Description of the Research**

You are invited to participate in a research study conducted by the National University of Medical Sciences (NUMS). The purpose of this study is to investigate the association of Surfactant Protein A (SP-A) and Surfactant Protein D (SP-D) gene polymorphisms with active tuberculosis and latent tuberculosis infection in the Pakistani population. The study aims to evaluate whether variations in these genes influence susceptibility, immune response, and disease progression through laboratory-based genetic analysis and clinical assessment.

**Procedures**

If you agree to participate, you will be asked to provide a blood sample and complete a structured questionnaire that includes relevant clinical and demographic information. The collected blood samples will be used for DNA extraction and genetic analysis to identify SP-A and SP-D gene polymorphisms.

**Risks and Discomforts**

There are no significant risks associated with participation in this study. Blood sample collection may cause minimal and temporary discomfort at the site of needle insertion.

**Potential Benefits**

Although there may be no direct benefit to you, this study may help in understanding the genetic factors associated with susceptibility to active and latent tuberculosis. The findings may contribute to improved risk assessment, early detection strategies, and the development of targeted prevention and treatment approaches.

**Protection of Confidentiality**

Your name or any identifying information will not be written on questionnaires or laboratory samples. All collected data will be coded and kept confidential, and access will be limited to authorized research personnel only.

**Voluntary Participation**

Your participation in this study is entirely voluntary. You may refuse to participate or withdraw from the study at any time without any penalty or effect on your medical care.

**Contact Information**

If you have any questions or concerns about this study, you may contact the research team at, 03237376736.

**Consent**
I have read and understood the information provided above. I have been allowed to ask questions, and all my questions have been answered satisfactorily. I voluntarily agree to participate in this study.

**Participant’s Signature:___________________________	Date: ___________________________**

***A copy of this consent form will be provided to you.***

**TB genetics Pakistan CRF: Active TB patients**

1. **PERSONAL DETAILS**

First name

Last name

Address

Mobile number

**Alternative contact:**

FAMILY MEMBER / FRIEND THAT MAY BE CONTACTED

Contact person/relationship

**/**

Contact person mobile number

2. **STUDY VISIT DETAILS AND ELIGIBILITY**

Date of visit

**Eligibility:**

| **Inclusion criteria** | **Yes** | **No** |
| --- | --- | --- |
| 1. Aged 16 or above |  |  |
| 2. Newly diagnosed active TB about to start treatment |  |  |
| 3. Gives written informed consent to participate in the study |  |  |
| **Exclusion criteria** |  |  |
| 1. Known Anaemia (Hb <10 g/dl) |  |  |
| 2. Known HIV infection |  |  |
| 3. Pregnant or breastfeeding |  |  |

**Note**:

* If the answer to any of the inclusion criteria is **NO**, the subject is not eligible for the study *

**If the answer to any of the exclusion criteria is **YES**, the subject is not eligible for the study**

3. **DEMOGRAPHIC AND CLINICAL DETAILS**

| Age |  |
| --- | --- |
| Gender |  |
| Ethnic origin |  |
| Site of disease |  |
| Basis of diagnosis (Smear, GeneXpert, other) |  |
| Duration of symptoms |  |
| Organism isolated |  |
| Drug-susceptibility |  |
| Previous anti-TB treatment |  |
| Details if so |  |
| Past medical history |  |
| Current medications |  |
| Smoking history | o Current ex-smoker never |
| BCG status (BCG scar present or absent?) |  |

4.
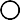

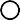
**RESULTS**

| **Laboratory test done** | **Result** |
| --- | --- |
| Haemoglobin level if available [state N/A if not available] |  |
| Chest x-ray if available [state N/A if not available]  Date: |  |

**Actual volumes (mL) of blood collected:**

| **Tube** | **Target volume** | **Actual volume** |
| --- | --- | --- |
| EDTA | 5 mL |  |

Date and time of blood collection: _

Date and time of receipt of blood in laboratory: _	_

Received by (name of laboratory personnel):

**Details of person completing CRF:**

Name	Signature _

**TB genetics Pakistan CRF: Healthy controls**

1. **PERSONAL DETAILS**

First name

Last name

Address

Mobile number

**Alternative contact:**

FAMILY MEMBER / FRIEND THAT MAY BE CONTACTED

Contact person/relationship

**/**

Contact person mobile number

2. **STUDY VISIT DETAILS AND ELIGIBILITY**

Date of visit

**Eligibility:**

| **Inclusion criteria** | **Yes** | **No** |
| --- | --- | --- |
| 1. Aged 16 or above |  |  |
| 2. Doesn’t have symptoms of TB |  |  |
| 3. Gives written informed consent to participate in the study |  |  |
| **Exclusion criteria** |  |  |
| 1. Is diagnosed with TB/ has been in contact with TB patient |  |  |
| 2. Known Anaemia (Hb <10 g/dl) |  |  |
| 3. Known HIV infection |  |  |
| 4. Pregnant or breastfeeding |  |  |

**Note**:

- If the answer to any of the inclusion criteria is **NO**, the subject is not eligible for the study *

**If the answer to any of the exclusion criteria is **YES**, the subject is not eligible for the study**

3. **DEMOGRAPHIC AND CLINICAL DETAILS**

| Age |  |
| --- | --- |
| Gender |  |
| Ethnic origin |  |
| Previous anti-TB treatment |  |
| Details if so |  |
| Past medical history |  |
| Current medications |  |
| Smoking history | o Current ex-smoker never |
| BCG status (BCG scar present or absent?) |  |

4.
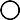

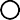
**RESULTS**

| **Laboratory test done** | **Result** |
| --- | --- |
| Haemoglobin level if available [state N/A if not available] |  |
| Chest x-ray if available [state N/A if not available]  Date: |  |

**Actual volumes (mL) of blood collected:**

| **Tube** | **Target volume** | **Actual volume** |
| --- | --- | --- |
| EDTA | 5 mL |  |

Date and time of blood collection: _

Date and time of receipt of blood in laboratory: _	_

Received by (name of laboratory personnel):

**Details of person completing CRF:**

Name	Signature _

**TB genetics Pakistan CRF: TB Contacts**

1. **PERSONAL DETAILS**

First name

Last name

Address

Mobile number Alternative contact:

FAMILY MEMBER / FRIEND THAT MAY BE CONTACTED

Contact person/relationship

**/**

Contact person mobile number

2. **STUDY VISIT DETAILS AND ELIGIBILITY**

Date of visit

**Eligibility:**

____	_	_

| **Inclusion criteria** | **Yes** | **No** |
| --- | --- | --- |
| 1. Aged 16 or above |  |  |
| 2. Household contact with index case of pulmonary TB within the last   6 months |  |  |
| 3. Asymptomatic (*i.e.,*no cough, weight loss, fever, night sweats,   lymphadenopathy or any other symptom / sign of active TB) |  |  |
| 4. Gives written informed consent to participate in the study |  |  |
| **Exclusion criteria** |  |  |
| 1. Clinical suspicion of active TB |  |  |
| 2. Known anaemia (Hb <10 g/dl) |  |  |
| 3. Pregnant or breastfeeding |  |  |
| 4. Known HIV infection |  |  |

**Note**:

- If the answer to any of the inclusion criteria is **NO**, the subject is not eligible for the study *

**If the answer to any of the exclusion criteria is **YES**, the subject is not eligible for the study**

3. **DEMOGRAPHIC AND CLINICAL DETAILS**

| Age |  |
| --- | --- |
| Gender |  |
| Ethnic origin |  |
| Occupation |  |
| Place of contact with index pulmonary TB case | - Household - Other |
| Index case’s relationship with the participant | - Parent/sibling - Other |
| Participant’s proximity to index case | - Sleep in the same bed - Sleep in different bed but same room - Sleeps in a different room, same house - Sleeps in a different house |
| Activities shared with the index case | - Every day, >=50% of the day - Every day, <50% of the day - Not every day |
| How long was the index case coughing before  their anti-TB treatment was started? |  |
| Past medical history |  |
| Current medications |  |
| Smoking history | o Current ex-smoker never |
| BCG status (BCG scar present or absent?) |  |

4.
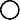

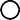
**RESULTS:**

| **Test done** | **Result** |
| --- | --- |
| Haemoglobin level if available [state N/A if not available] |  |

**Actual volumes (mL) of blood collected:**

| **Tube** | **Target volume** | **Actual volume** |
| --- | --- | --- |
| EDTA | 5 mL |  |

Date and time of blood collection: _	Date and time of receipt of blood in laboratory: _	_ Received by (name of laboratory personnel):

**Details of person completing CRF:**

Name	Signature _

**Agarose Gel Pictures**

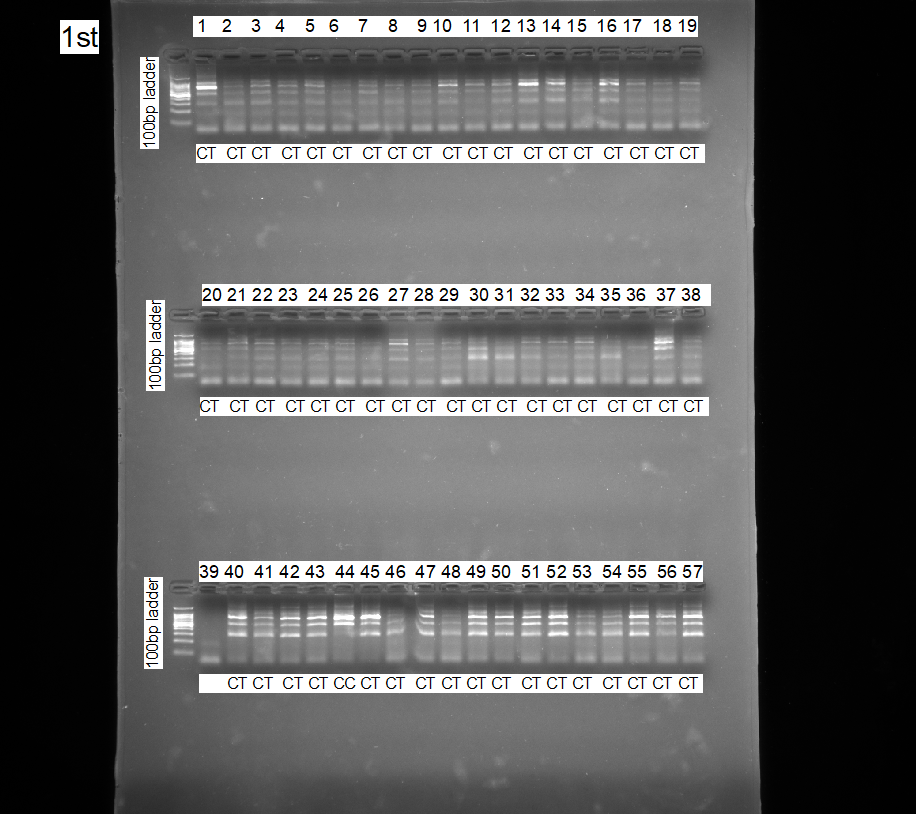

**Agarose gel pictures representing genotype pattern for rs1059047 using ARMs PCR**

Lane L represents 100 bp DNA ladder. The other lanes (1-57) refer to genotype pattern of TB patients (TP). Genotypes are characterized as 663 for all genotypes, 663+450 for CC, 663+264 for TT, and 663+450+264 CT.

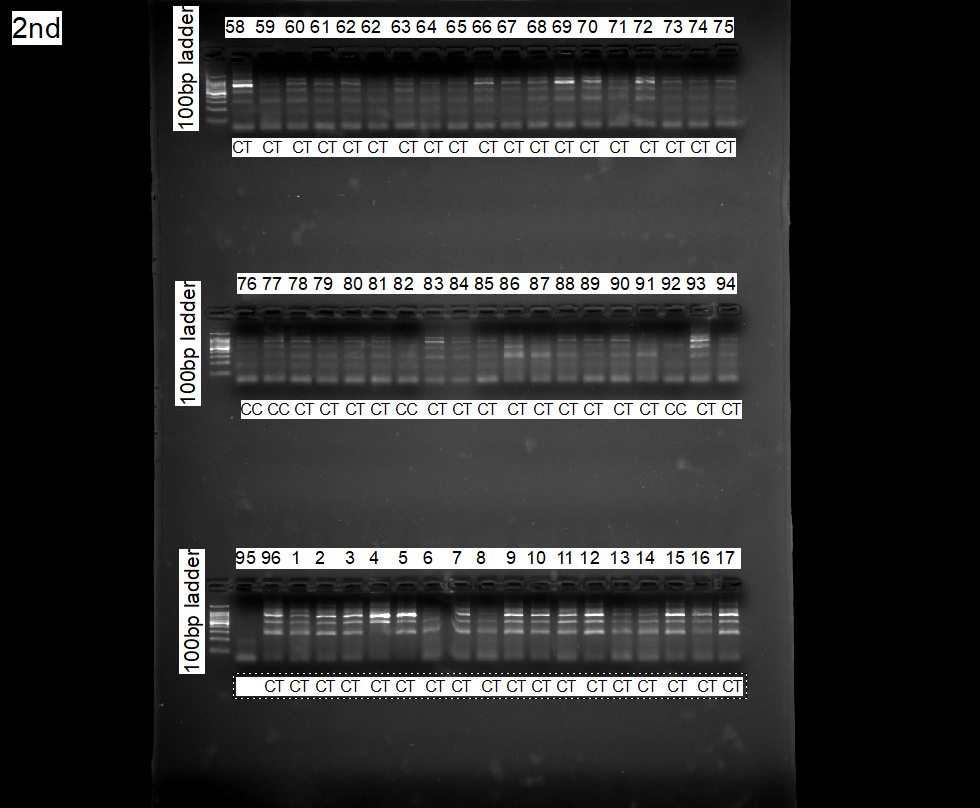

**Agarose gel pictures representing genotype pattern for rs1059047 using ARMs PCR**

Lane L represents 100 bp DNA ladder. The other lanes (TP57-96 and HC 1-17) refer to genotype pattern of TB patients (TP). Genotypes are characterized as 663 for all genotypes, 663+450 for CC, 663+264 for TT, and 663+450+264 CT.

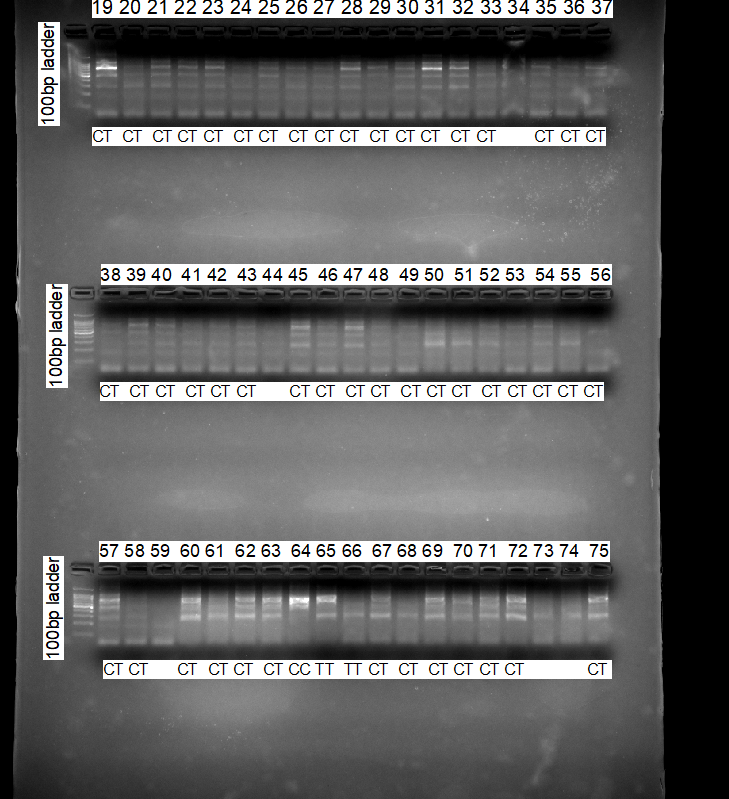

**Agarose gel pictures representing genotype pattern for rs1059047 using ARMs PCR**

Lane L represents 100 bp DNA ladder. The other lanes (HC 19-75) refer to genotype pattern of TB patients (TP). Genotypes are characterized as 663 for all genotypes, 663+450 for CC, 663+264 for TT, and 663+450+264 CT.

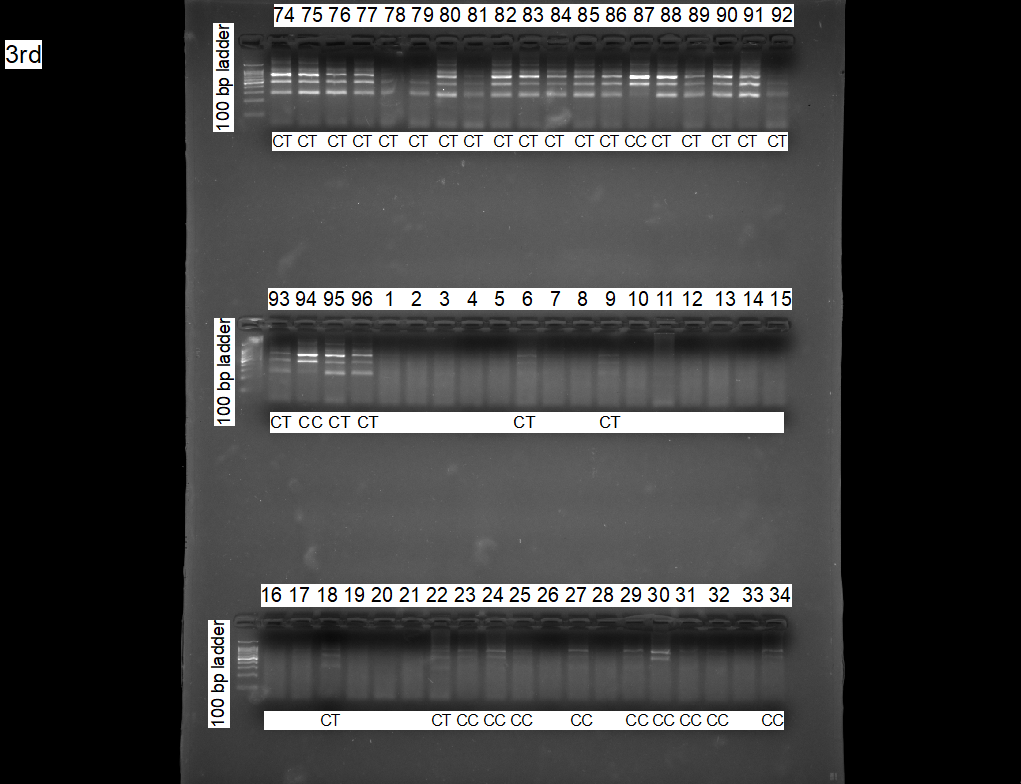

**Agarose gel pictures representing genotype pattern for rs1059047 using ARMs PCR**

Lane L represents 100 bp DNA ladder. The other lanes (HC 74-96 and TC 1-34) refer to genotype pattern of TB patients (TP). Genotypes are characterized as 663 for all genotypes, 663+450 for CC, 663+264 for TT, and 663+450+264 CT.

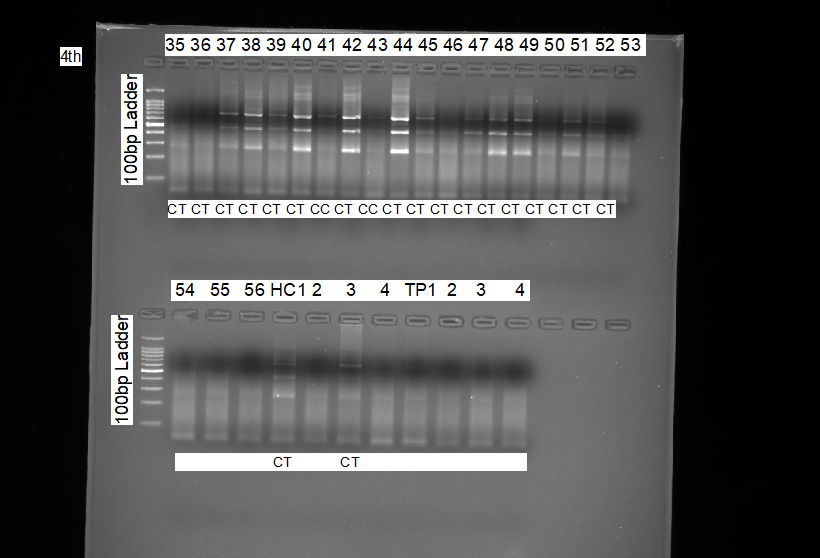

TC 35-56

**Agarose gel pictures representing genotype pattern for rs1059047 using ARMs PCR**

Lane L represents 100 bp DNA ladder. The other lanes (TC 35-56) refer to genotype pattern of TB patients (TP). Genotypes are characterized as 663 for all genotypes, 663+450 for CC, 663+264 for TT, and 663+450+264 CT.

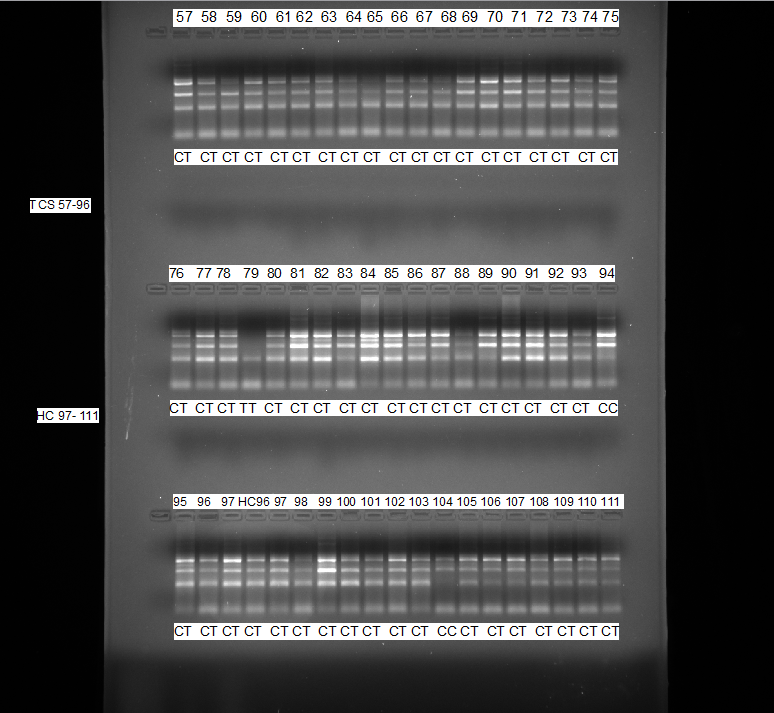

**Agarose gel pictures representing genotype pattern for rs1059047 using ARMs PCR**

Lane L represents 100 bp DNA ladder. The other lanes (TC57-96 and HC 96-111) refer to genotype pattern of TB patients (TP). Genotypes are characterized as 663 for all genotypes, 663+450 for CC, 663+264 for TT, and 663+450+264 CT.

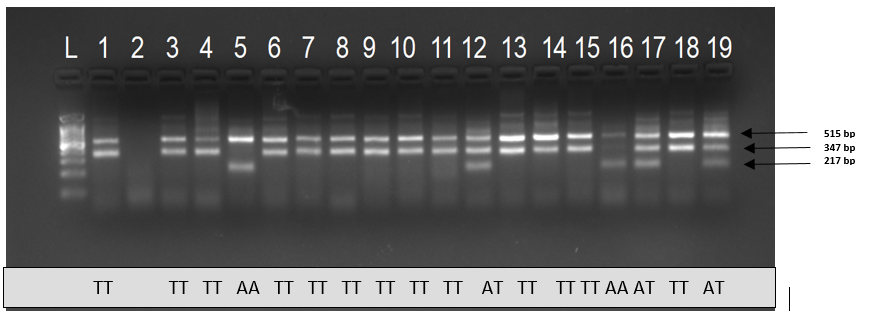

Agarose gel picture representing genotype pattern for SNP rs3088308 using ARMS-PCR. Lane L represents 100 bp DNA ladder. The other lanes (1-19) refer to genotype pattern of Healthy controls. Genotypes are characterized as TT (515 bp, 347 bp), AA (515 bp and 217 bp) and AT (515 bp, 347 bp and 217 bp).

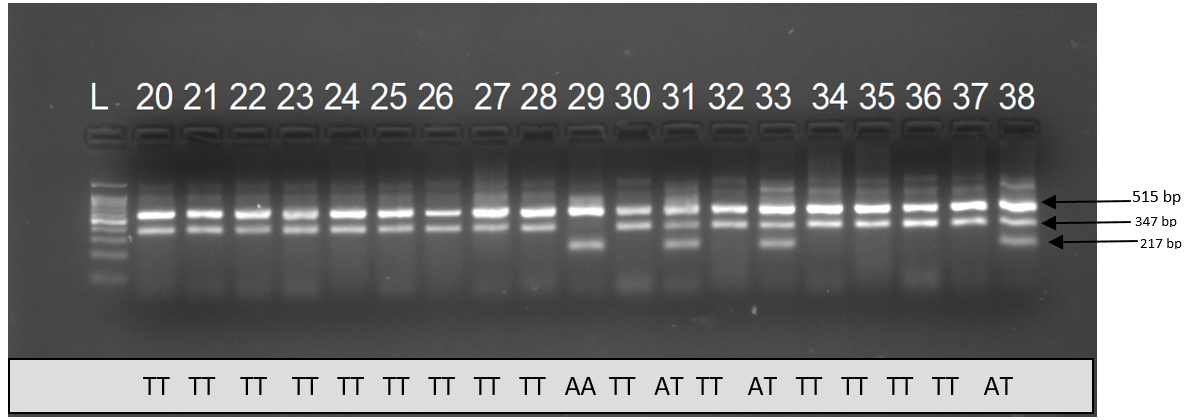

Agarose gel picture representing genotype pattern for SNP rs3088308 using ARMS-PCR. Lane L represents 100 bp DNA ladder. The other lanes (20-38) refer to genotype pattern of Healthy controls. Genotypes are characterized as TT (515 bp, 347 bp), AA (515 bp and 217 bp) and AT (515 bp, 347 bp and 217 bp).

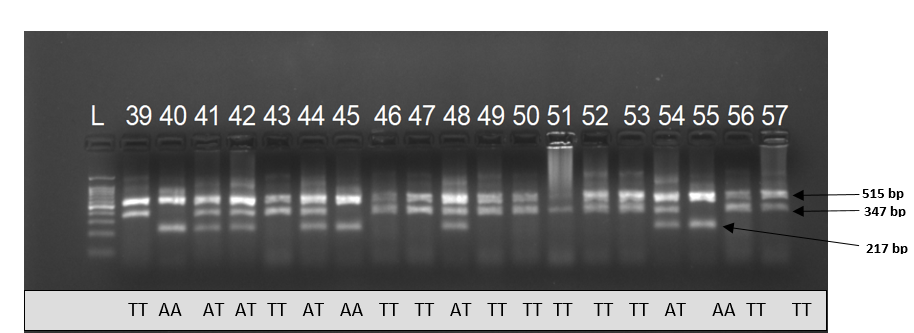
 Agarose gel picture representing genotype pattern for SNP rs3088308 using ARMS-PCR. Lane L represents 100 bp DNA ladder. The other lanes (39-57) refer to genotype pattern of Healthy controls. Genotypes are characterized as TT (515 bp, 347 bp), AA (515 bp and 217 bp) and AT (515 bp, 347 bp and 217 bp).

**`**

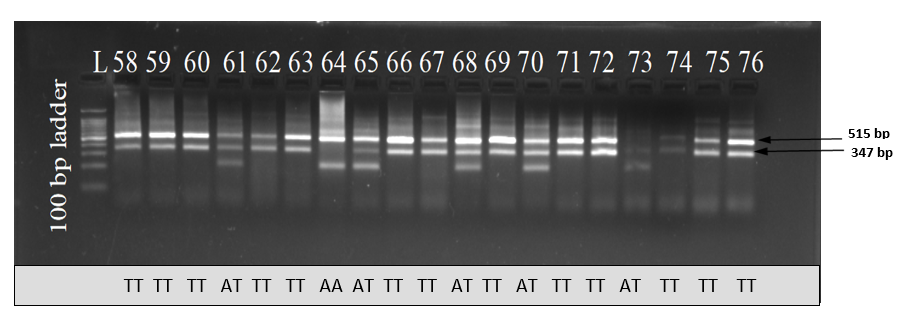
 Agarose gel picture representing genotype pattern for SNP rs3088308 using ARMS-PCR. Lane L represents 100 bp DNA ladder. The other lanes (58-76) refer to genotype pattern of Healthy controls. Genotypes are characterized as TT (515 bp, 347 bp), AA (515 bp and 217 bp) and AT (515 bp, 347 bp and 217 bp).

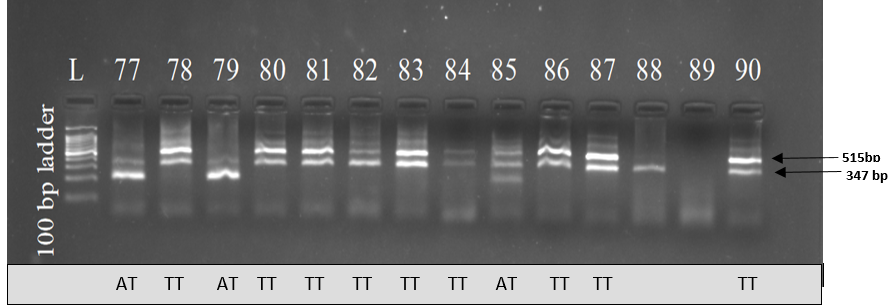
 Agarose gel picture representing genotype pattern for SNP rs3088308 using ARMS-PCR. Lane L represents 100 bp DNA ladder. The other lanes (77-90) refer to genotype pattern of Healthy controls. Genotypes are characterized as TT (515 bp, 347 bp), AA (515 bp and 217 bp) and AT (515 bp, 347 bp and 217 bp).

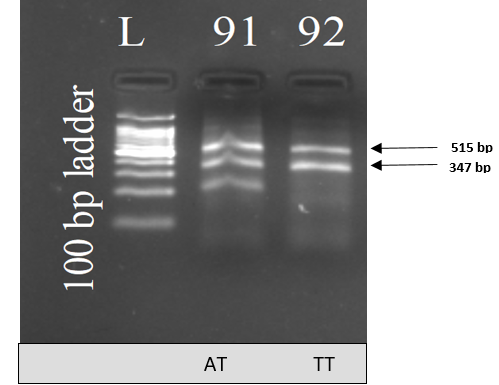

Agarose gel picture representing genotype pattern for SNP rs3088308 using ARMS-PCR. Lane L represents 100 bp DNA ladder. The other lanes (1-19) refer to genotype pattern of Healthy controls. Genotypes are characterized as TT (515 bp, 347 bp), AA (515 bp and 217 bp) and AT (515 bp, 347 bp and 217 bp).

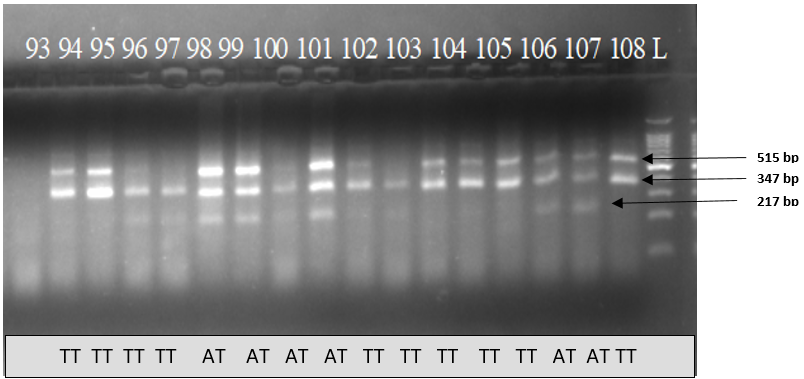

Agarose gel picture representing genotype pattern for SNP rs3088308 using ARMS-PCR. Lane L represents 100 bp DNA ladder. The other lanes (93-108) refer to genotype pattern of Healthy controls. Genotypes are characterized as TT (515 bp, 347 bp), AA (515 bp and 217 bp) and AT (515 bp, 347 bp and 217 bp).

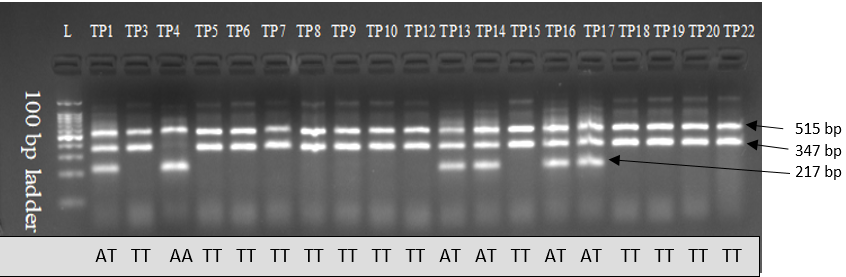

Agarose gel picture representing genotype pattern for SNP rs3088308 using ARMS-PCR. Lane L represents 100 bp DNA ladder. The other lanes (TP1-A22) refer to genotype pattern of TB patients. Genotypes are characterized as TT (515 bp, 347 bp), AA (515 bp and 217 bp) and AT (515 bp, 347 bp and 217 bp).

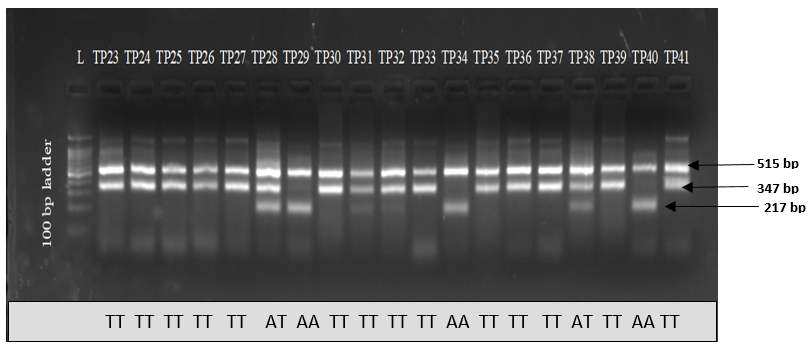

Agarose gel picture representing genotype pattern for SNP rs3088308 using ARMS-PCR. Lane L represents 100 bp DNA ladder. The other lanes (TP23-A41) refer to genotype pattern of TB patients. Genotypes are characterized as TT (515 bp, 347 bp), AA (515 bp and 217 bp) and AT (515 bp, 347 bp and 217 bp).

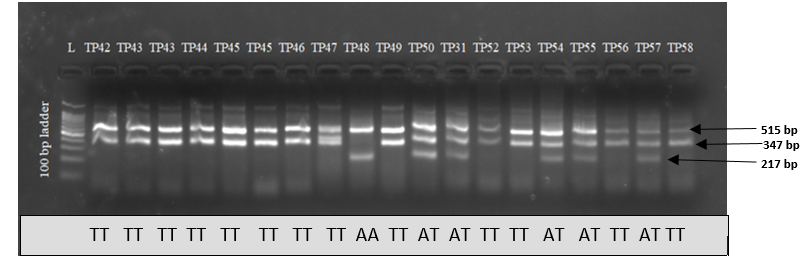
 Agarose gel picture representing genotype pattern for SNP rs3088308 using ARMS-PCR. Lane L represents 100 bp DNA ladder. The other lanes (TP42-TP58) refer to genotype pattern of TB patients. Genotypes are characterized as TT (515 bp, 347 bp), AA (515 bp and 217 bp) and AT (515 bp, 347 bp and 217 bp).

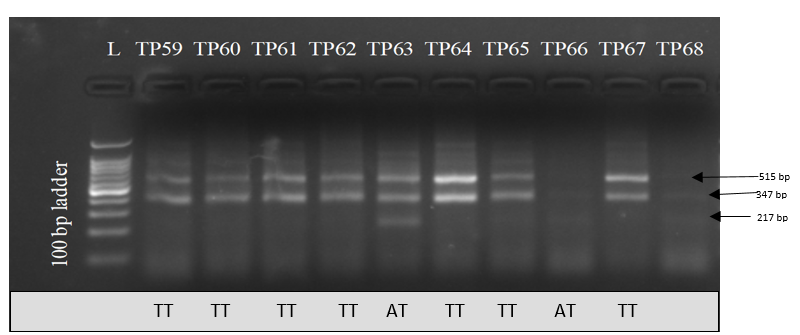
 Agarose gel picture representing genotype pattern for SNP rs3088308 using ARMS-PCR. Lane L represents 100 bp DNA ladder. The other lanes (TP59-TP68) refer to genotype pattern of TB patients. Genotypes are characterized as TT (515 bp, 347 bp), AA (515 bp and 217 bp) and AT (515 bp, 347 bp and 217 bp).

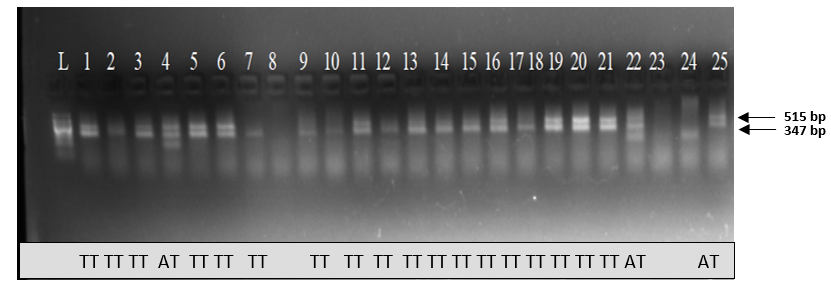
 Agarose gel picture representing genotype pattern for SNP rs3088308 using ARMS-PCR.Lane L represents 100 bp DNA ladder. The other lanes (1-25) refer to genotype pattern of TB Contacts. Genotypes are characterized as TT (515 bp, 347 bp), AA (515 bp and 217 bp) and AT (515 bp, 347 bp and 217 bp).

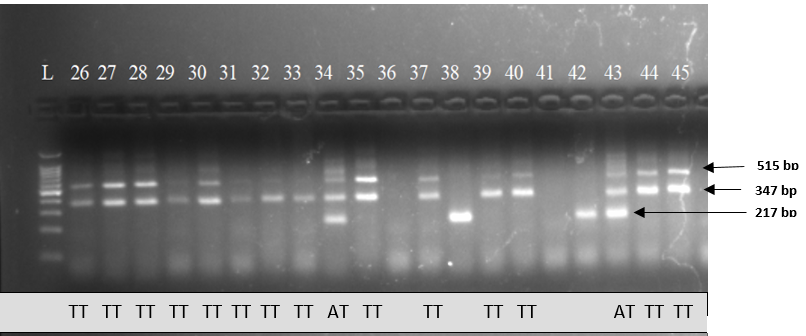
Agarose gel picture representing genotype pattern for SNP rs3088308 using ARMS-PCR. Lane L represents 100 bp DNA ladder. The other lanes (26-45) refer to genotype pattern of TB Contacts. Genotypes are characterized as TT (515 bp, 347 bp), AA (515 bp and 217 bp) and AT (515 bp, 347 bp and 217 bp).

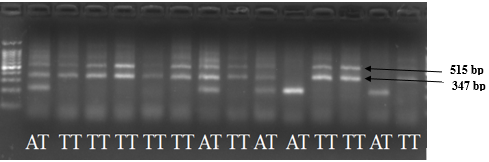

Agarose gel picture representing genotype pattern of TB contacts for SNP rs3088308 using ARMS-PCR. Genotypes are characterized as TT (515 bp, 347 bp), AA (515 bp and 217 bp) and AT (515 bp, 347 bp and 217 bp).
